# The Fractional Epidemics Theory: Managing and Ending an Epidemic

**DOI:** 10.1101/2020.11.08.20227793

**Authors:** T. Duclos, T. Reichert

## Abstract

Susceptible–infectious–recovered (SIR) models are widely used for estimating the dynamics of epidemics and project that social distancing “flattens the curve”, i.e., reduces but delays the peak in daily infections, causing a longer epidemic. Based on these projections, individuals and governments have advocated lifting containment measures such as social distancing to shift the peak forward and limit societal and economic disruption. Paradoxically, the COVID-19 pandemic data exhibits phenomenology opposite to the SIR models’ projections. Here, we present a new model that replicates the observed phenomenology and quantitates pandemic dynamics with simple and actionable analytical tools for policy makers. Specifically, it offers a prescription of achievable and economically palatable measures for ending an epidemic.

**One Sentence Summary:** The SIR epidemic models are wrong; a new model offers achievable and economically viable measures for ending an epidemic.

## Main Text

The susceptible–infectious–recovered (SIR) models *(1)*, widely used in predictions of epidemic spread, predict that social distancing will “flatten the curve”, i.e., reduce the total number of cases and delay the peak of new daily cases. The peak in distress is reduced by a modest fraction; but the misery lengthens in approximately inverse proportion. With this image highlighted in popular media, concerns about the economic devastation projected to be associated with such measures applied in response to COVID-19 has many individuals and governments advocating that social distancing measures be lifted early *(2)* to shift the peak forward and end the economic, educational, and social disruption sooner.

In the real world, countries have applied different mitigation measures *(3-5)* to control the COVID-19 pandemic, with varied success. These natural experiments have provided an opportunity to compare model projections of the effect of different containment measures against real world data.

### Testing the SIR Model

A review of the seminal paper for SIR models reveals a fundamental flaw early in the model formulation. The paper states that “…the chance of an infection is proportional to the number infected on one hand, and to the number not yet infected on the other” *(1, p. 703)*. The authors argue that the change in the number of infections at any time is proportional to the number of people infected, multiplied by the number of people not yet infected. Though seemingly logical, the statement implies that people who have not yet been infected associate only with infected people and vice versa. However, the interactions between a recovered person and a susceptible or infected person must also be accounted for because they “use up” interactions that could have occurred between an infected person and a susceptible person. Due to the omission of these “unsuccessful” interactions in the model formulations, the equations that describe the change in infected, recovered and susceptible people (Equations 1, 2 and 3) in the SIR model and its variants lack any terms related to the recoveries.

One consequence of this omission can be seen in Equation 3 where the change in the recovered population as a function of time is depicted as linearly related to the total size of the infected population alone. This cannot be correct because the members of the infectious population that recover at any specific time were infected at a time in the past separated from the current time by an interval called the recovery time. Given the various nonlinear relationships embedded in the model, it is illogical that the change in the recovered population, would be directly proportional to the total size of the currently infected population.

To determine the degree to which these flaws affect the model performance, we tested the capability of an SIR model *(6)* to predict observed trends in the COVID pandemic using the following equations:

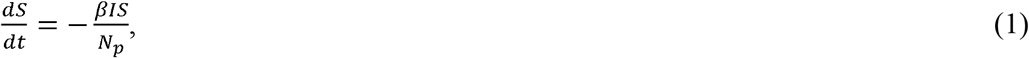

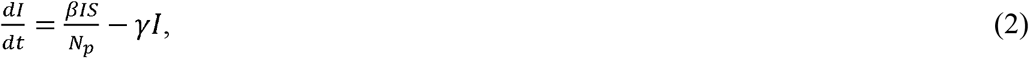

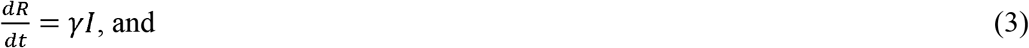

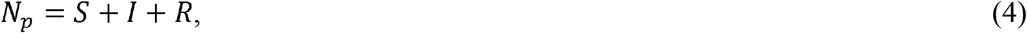

where *S* = number of people susceptible to infection at time *t*; *I* = number of people infected at time *t*; *R* = number of people recovered at time *t*; *N*_*p*_ = total number of people in the population; *β* = rate of contact and transmission; *t*_*r*_ = time of infectiousness; and *γ* = rate of recoveries = 1/*t*_*r*_.

In Figure 1A, we see that the SIR model projects that the time at which the total number of cases levels off is delayed as social distancing increases (represented by decreasing *β*), while the ultimate total case number remains quite similar. Increasing social distancing also delays and reduces (here by about half) the peak of new infections, while resulting in a broader peak (Fig. 1B).

**Fig. 1.**
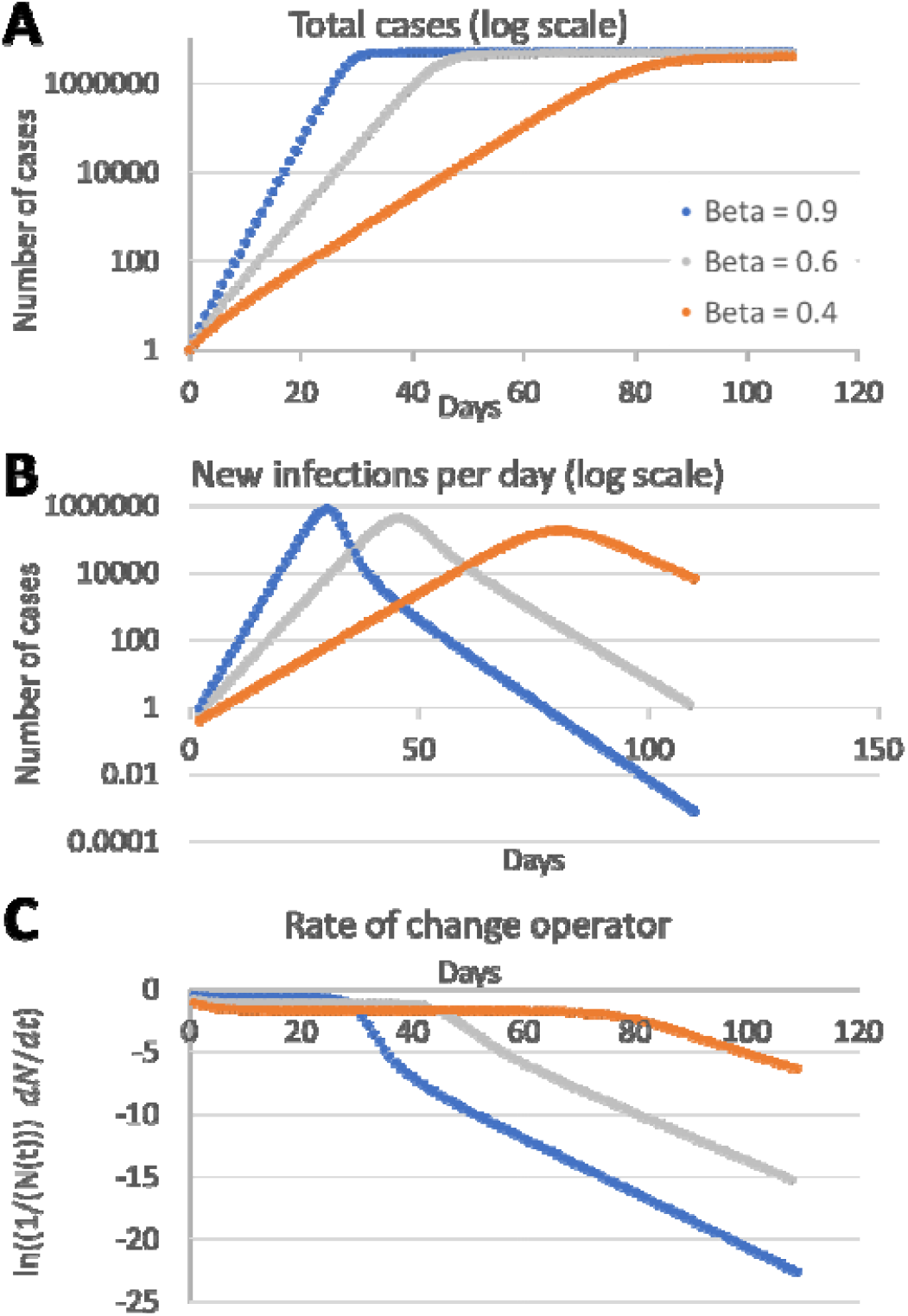
SIR model predictions. Rate of contact and transmission (β) decreases with lower social interaction. Rate of recoveries (*γ*) = 0.2 for all plots. As β decreases, the daily total of cases increases slower and plateaus later (**A**), daily new infections project to later peaks (**B**), and the slope of the RCO operator steepens progressively later (**C**).

To further enhance the analysis of the model, we introduce here a new quantity called the rate of change operator (RCO, Fig. 1C); the logarithm of the ratio of the change in daily cases to the time-associated total of cases *(6.)* This formulation produces a new function that summarizes epidemic dynamics. Applied to the SIR model projections, initial flat segments in the graphs reflect near-exponential growth and each flat line changes abruptly near the peak in daily cases, with the time to the change proportional to the strength of containment. The slope of the new line segment is inversely proportional to the strength of containment.

As a test of the SIR projections, we compared case data between Sweden and New Zealand (Fig. 2A, C, E), and between South Korea and Italy (Fig. 2B, D, F), pairs of countries that have comparable population densities but that implemented mitigation measures with different timings and intensities *(3-5, 8)*. In particular, New Zealand and South Korea had much earlier and stronger social distancing measures than Italy and Sweden.

**Fig. 2.**
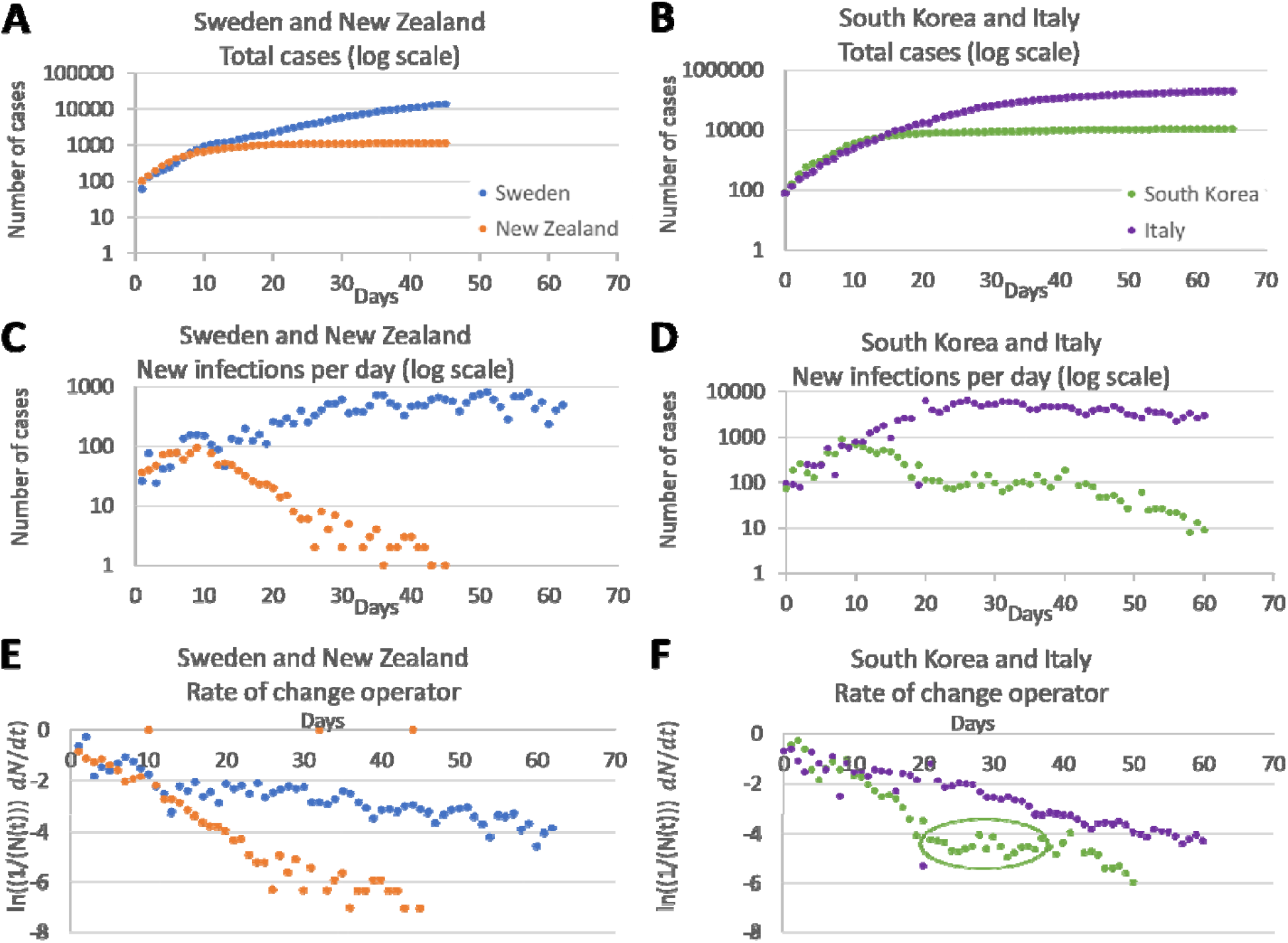
Data reported from different countries during the COVID-19 pandemic. Reported data from pairs of countries (Sweden, blue, and New Zealand, orange; and South Korea, green, and Italy, purple), referenced to a day when each pair had nearly equal numbers of new cases. (**A, B**) Daily total infections. (**C, D**) Daily new infections. (**E, F**) RCO plots of daily new infections. A new outbreak occurred around day 25 in South Korea *(3)* and created a plateau in the RCO curve (circled). All trends shown in this figure are the opposite of the trends in Figure 1.

In contrast to the SIR model predictions (Fig. 1), the country data (Fig. 2) show that stronger social distancing measures are associated with an earlier and lower peak in new infections, an earlier levelling off at a lower number of total cases, and a steeper RCO slope. All three trends in Fig. 2 demonstrate that the SIR model (Fig 1A-C) is not merely inaccurate but actually projects epidemic data to trend in the opposite direction to the reported data. The SIR model simply does not match real world experience. All variants of the SIR model suffer from this fault and predict that the peak in daily cases will be delayed as social distancing increases.

Because the faults in the SIR models stem from their mathematical foundations, no amount of curve fitting will enable them to predict the observed trends. Rather, a new model formulation, built on the proper foundation, is needed.

### The Fractional Epidemic Model

Taking a new approach, we developed a new model using basic principles incorporating the proper population interactions *(6)*. Because the model was developed using insight gained by casting the number of infected and recovered individuals as fractions of the total case count, we call the model the fractional epidemic (FE) model.

The model is described by the following equations:

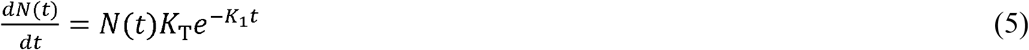

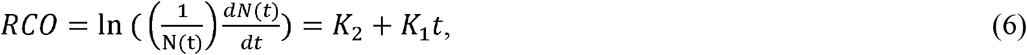

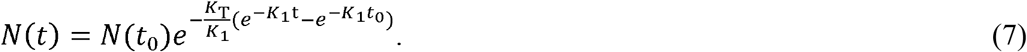

*I*(*t*) represents the currently infectious people, *R*(*t*) people who have been infected but are no longer contagious, and *N*(*t*) = *R*(*t*) + *I*(*t*). *K*_*T*_ is a property of the disease that is a constant representing disease transmissibility, *P*_*c*_ is the instantaneous number of contacts each individual in *N(t)* has with the entire population, and 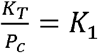 (therefore, *K*_1_ represents the behavior of the population). *t*_*0*_ is the time of the first data point of the model; if the model starts at the beginning of the epidemic, *t*_*0*_ = 0.

Figure 3A-C depicts the qualitative behavior of the FE model. With increased social distancing (higher *K*_*1*_), the total number of daily cases is lower and attained earlier (Fig. 3A), the peak in new cases is lower and occurs earlier (Fig. 3B), and the slope of the RCO curve is steeper (Fig. 3C).

**Fig. 3.**
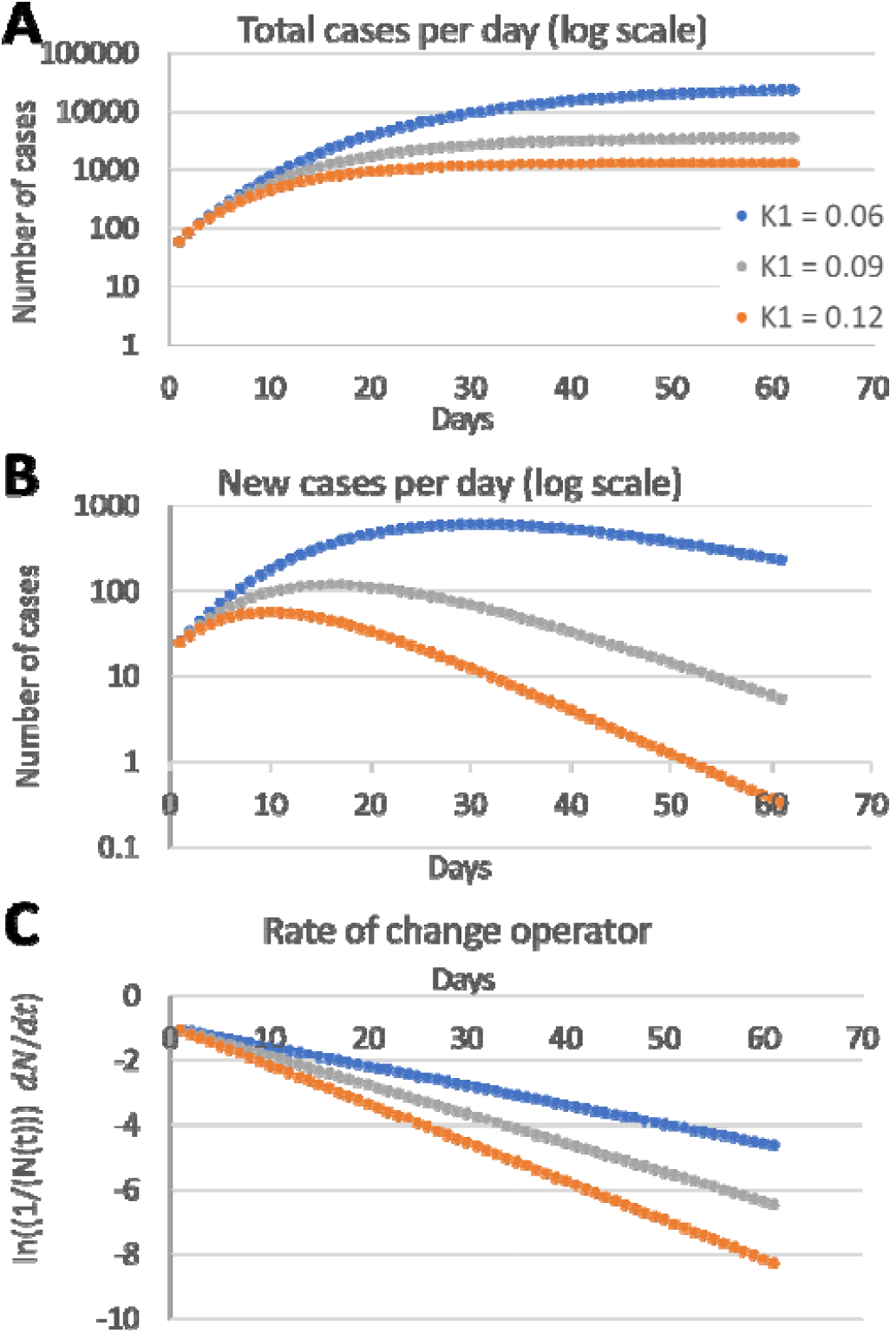
Fractional epidemic (FE) model predictions. (**A**) Projected trends of daily total cases (Equation 7). (**B**) Projected trends of daily new cases (Equation 5). (**C**) Rate of change (Equation 6). *K*_*2*_ = 1 for all plots. The FE model projects trends identical to those in the country-reported data (Fig. 2).

Unlike the SIR model trends (Figure 1A-C), the FE model trends (Figure 3A-C) are identical to those exhibited by the real-life COVID-19 data in Figure 2. Whereas the SIR models fail to predict the proper impact of social distancing, the FE model predicts the trends seen in the real data and, in particular, predicts that stronger social distancing produces an earlier peak of daily cases.

As a demonstration of the predictive capability of the FE model, we tested the model quantitatively on data from several countries by first estimating two country-specific constants, *K*_*1*_ and *K*_*2*_. These were found by using the RCO expression (Equation 6) to transform each country’s daily total cases to a new time series.

Shortly after intervention measures were imposed in each country, the resulting RCO time series (Fig. 4) have distinctive segments where the curves appear to become straight lines. The parameters *K*_*1*_ and *K*_*2*_, tabulated in in Table 1, were then estimated by fitting a linear expression to a small, early (nine data point) portion of these straight segments. The date ranges for these nine data point portions are listed in the table and these data points are highlighted in blue in Figures 4-6. The derived values for *K*_*1*_ and *K*_*2*_ were then used to predict the balance of the epidemic data that followed (grey dots in Figs. 5 and 6).

**Table 1.** Social containment parameters used to model total cases and new daily cases of infection.

| Country | $K_1$ | $K_2$ | $N_{(t_0)}$ | Date range for data fit |
| --- | --- | --- | --- | --- |
| South Korea | 0.195 | 1.623 | 3,526 | March 1 – March 9 |
| USA | 0.078 | 1.511 | 46,442 | March 26 – April 3 |
| Sweden | 0.061 | 2.297 | 4435 | April 1 – April 9 |
| Italy | 0.075 | 1.856 | 27,980 | March 17 – March 25 |
| Spain | 0.100 | 1.940 | 56,188 | March 27 – April 4 |
| New Zealand | 0.214 | 2.079 | 647 | 1 April 1 – April 9 |

**Fig. 4.**
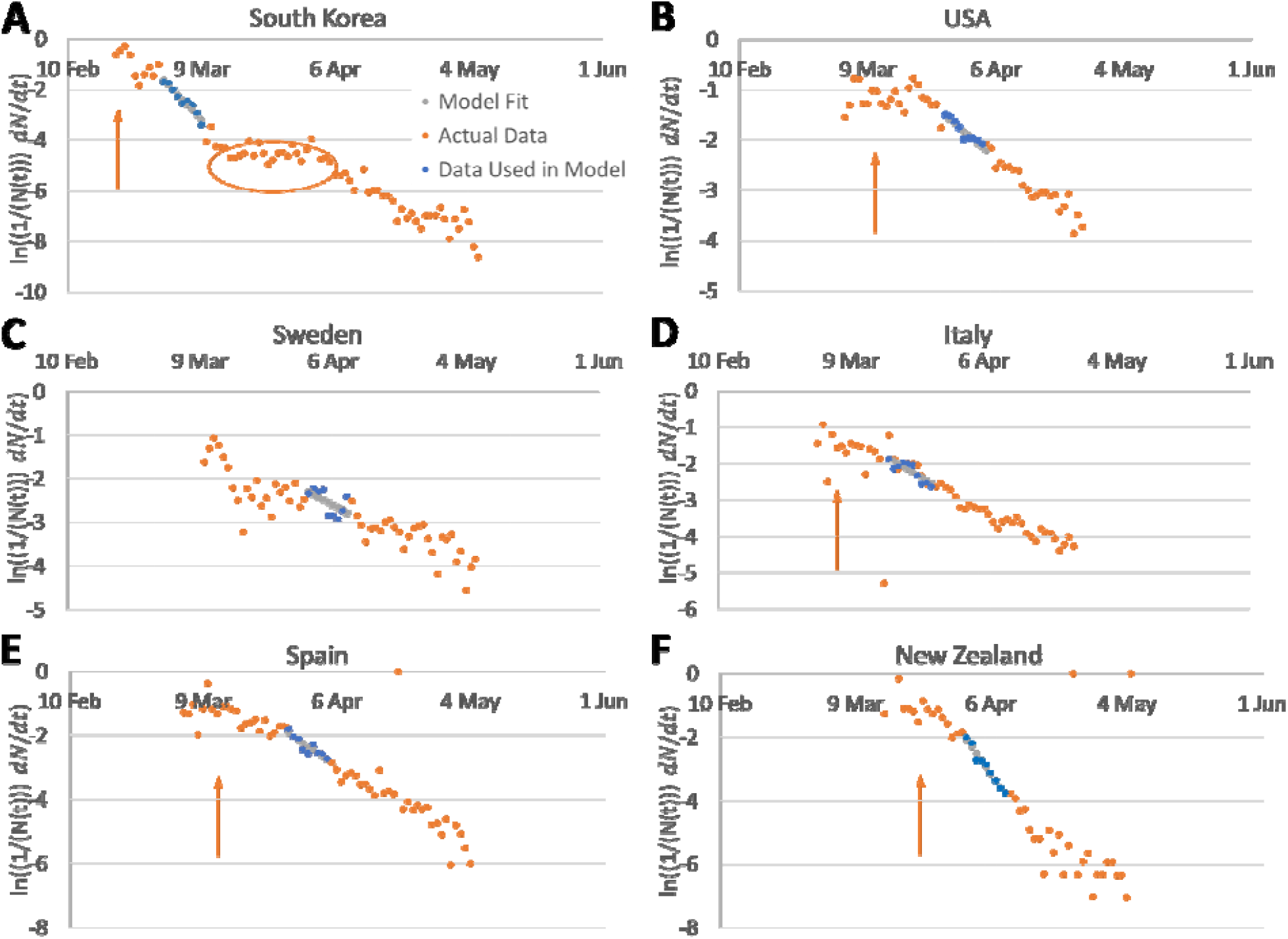
Rate of change operator (RCO) curves for the number of COVID-19 cases in various countries. An epidemic can be described by a piecewise linear model using the RCO (Equation 6). RCO curves change markedly, soon after containment measures are implemented (arrows: South Korea, February 21; USA, March 16; Italy, March 8–21; Spain, March 14; New Zealand, March 25. Sweden did not implement any specific containment measures).

**Fig. 5.**
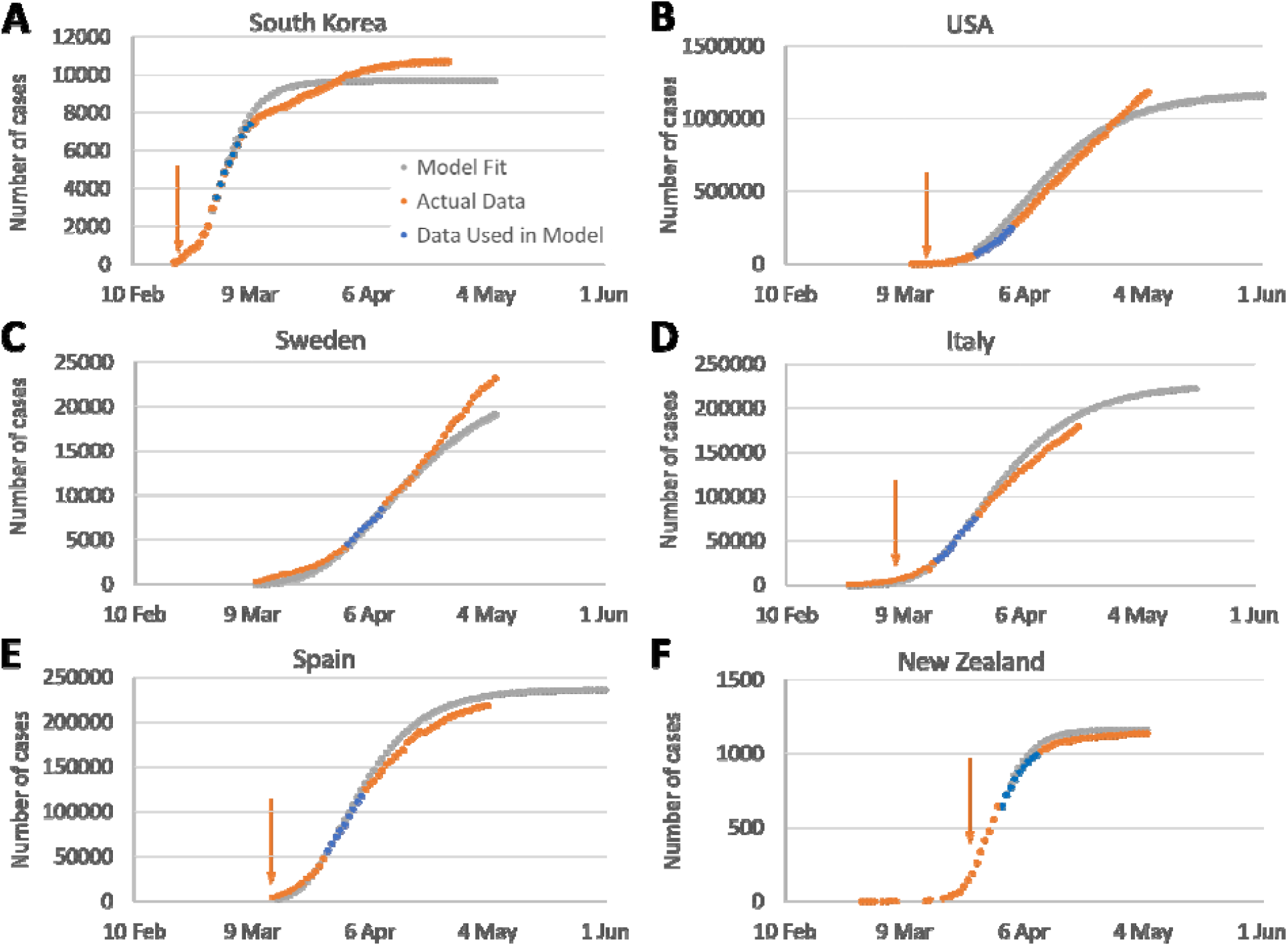
Fractional epidemic (FE) model predictions for daily total case counts. The blue data points in Figure 4 were used for the parameter fits. *R*^*2*^ > 0.97 for all countries except South Korea (*R*^*2*^ = 0.80). The FE model was calibrated using data from the date ranges listed in Table 1. These began 8–12 days after containment measures were imposed in 2020 (arrow in each panel: South Korea, February 21; USA, March 16; Italy, March 8; Spain, March 15; and New Zealand, March 25). Sweden did not implement any specific containment measures so the model calibration was begun on April 1, when the slope of the rate of change operator (RCO) curve first became steady.

**Fig. 6.**
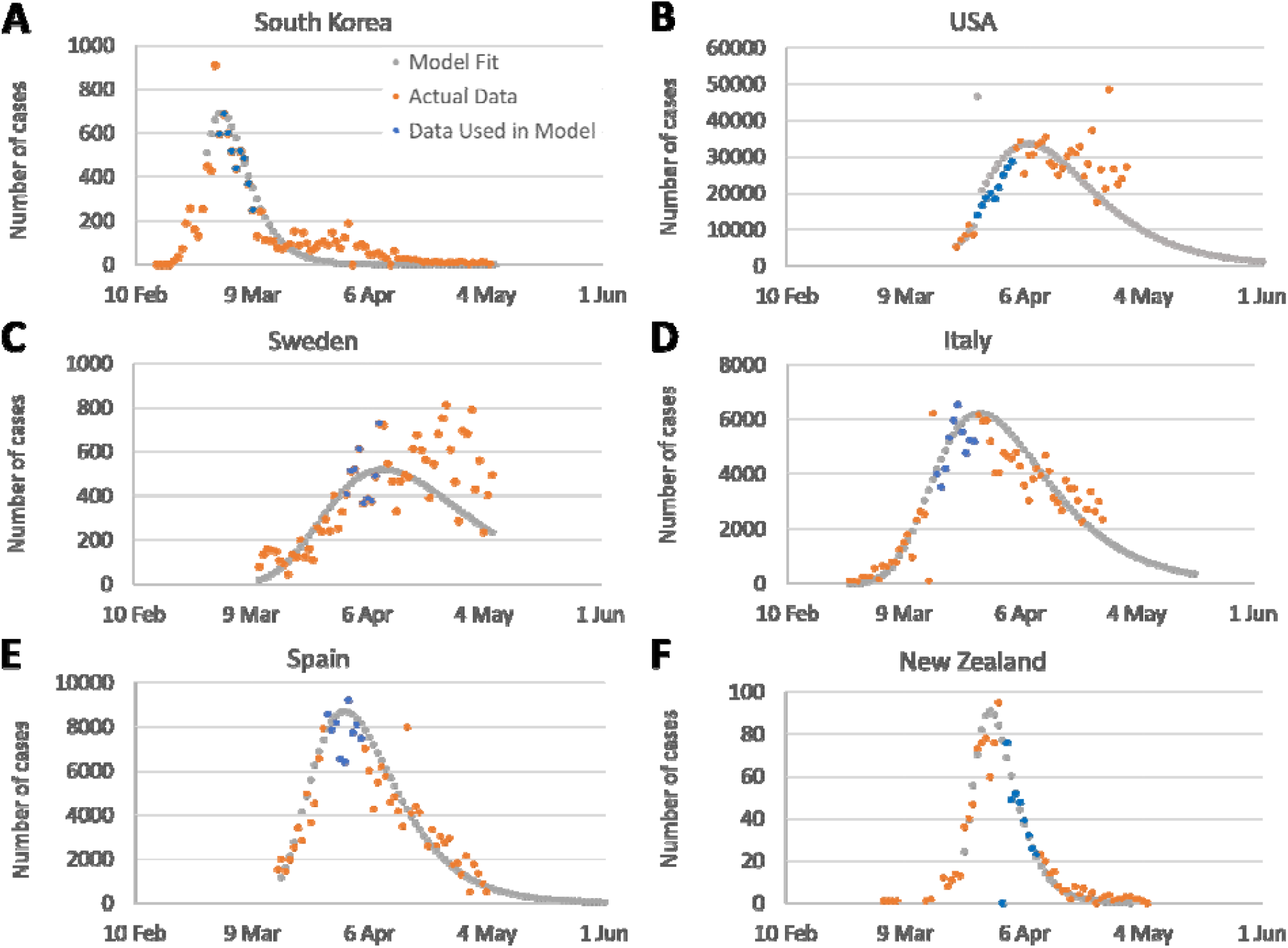
Fractional epidemic (FE) model predictions for number of new daily cases. (**A**) South Korea; (**B**) USA; (**C**) Sweden; (**D**) Italy; (**E**) Spain; and (**F**) New Zealand. R^2^ = 0.61–0.79 for the fit of the model to the data across countries.

Using this approach, the FE model correctly predicted the total cases with an *R*^*2*^ > 0.97 (Fig. 5, grey dots) for all but one of six countries for the 3 weeks following the 9-day data segment used to determine the two constants in the model (Table 1). The model also predicted daily new cases (Fig. 6) for these countries with an *R*^*2*^ range of 0.61–0.79 and the projected peak of new cases (Fig. 6, grey dots) was very close to the observed peak for all countries.

The FE model projections in Figures 5 and 6 are *not* fits to the full-length of the data shown. Rather, a short portion of the epidemic data, with specified characteristics, was excised for the purposes of determining coefficients of the model that were then used to determine the rest of the curves.

In an additional demonstration of the FE model’s veracity, we tested the assumption that *K*_*T*_ is a property of the disease; and therefore, should be the same for each country. Equation 71 *(6)* shows that the model parameters, expressed in a function, *f(N(t))*, should be linearly proportional to time with a constant of proportionality −*K*_*T*_. The fit of Equation 71 to the country data from Table 2 has an *R*^*2*^ = 0.95 and a slope (which is equal to −*K*_*T*_) of 0.19. This excellent result, along with Figs. 3, 5 and 6, demonstrates that the FE model correctly characterizes epidemic dynamics from multiple countries in a unified way; something SIR models simply cannot duplicate.

**Table 2.** Initial COVID pandemic data and social interaction parameters for various countries

| | Date of first case reported | Date of cases in calculation | Days | Cases on calculation date | Population density (people/km<sup>2</sup>) | $K_T$ | $K_{2b}$ | $K_{1b}$ | $N_\infty$ |
| --- | --- | --- | --- | --- | --- | --- | --- | --- | --- |
| South Korea | 20 Jan | 21 Feb | 32 | 155 | 508 | 0.19 | 1.7 | 4.99E-04 | 2.54E+165 |
| USA | 21 Jan | 19 Mar | 58 | 9415 | 37 | 0.19 | 1.7 | 6.90E-03 | 9.12E+11 |
| Sweden | 1 Feb | 7 Mar | 35 | 137 | 18 | 0.19 | 1.7 | 1.44E-02 | 5.32E+05 |
| Germany | 28 Jan | 19 Mar | 51 | 8198 | 232 | 0.19 | 1.7 | 1.09E-03 | 2.82E+75 |
| Italy | 31 Jan | 24 Feb | 24 | 132 | 208 | 0.19 | 1.7 | 1.22E-03 | 6.55E+67 |
| Spain | 1 Feb | 13 Mar | 41 | 3004 | 89 | 0.19 | 1.7 | 2.83E-03 | 1.37E+29 |
| New Zealand | 28 Feb | 19 Mar | 20 | 28 | 18 | 0.19 | 1.7 | 1.38E-02 | 9.55E+05 |
| France | 25 Jan | 13 Mar | 48 | 4499 | 122 | 0.19 | 1.7 | 2.08E-03 | 5.47E+39 |

$K_{1b}$ = baseline value of the social interaction parameter $K_1 = \frac{K_T A}{A_1 N_p}$ ; $\frac{N_p}{A}$ = population density; $A_l = 0.75$ found using Equation 67 (6); $K_{2b} = -\ln(K_T)$ ; $K_T$ , is a constant representing transmissibility of the disease; $N_\infty = e^{\frac{K_T}{K_{1b}}}$ , final number of cases if no containment is imposed. All dates are in 2020.

To illustrate why the FE model predicts epidemic dynamics while SIR models do not, we cast the FE model in terms of S, I and R equations:

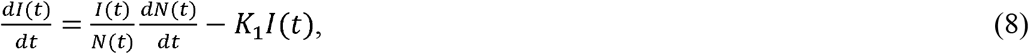

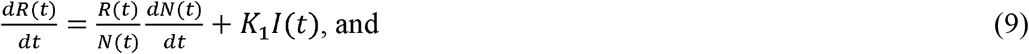

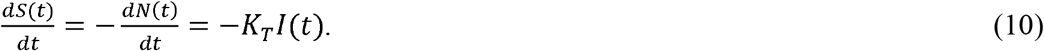

Additional insight is gained by using equation 22 *(6)*, to rewrite Equation 8:

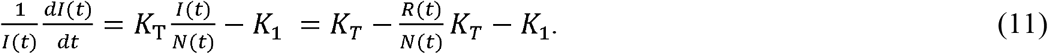

The term 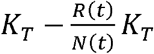 on the right hand side of Equation 11 describes the rate of successful infections per infectious individual and 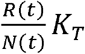 is the effect of the recovered population’s interactions on the infection rate. The ratio of the recovered population to the total number of cases is the rate transmission slows; and, coupled with the behavior of the population represented by *K*_*1*_, this controls the epidemic dynamics.

Because the recovered population interactions appear on the righthand side of Equations 8-11 either explicitly or implicitly (via 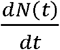), the FE model correctly projects the observed data from multiple countries in a unified way. In contrast, no terms for the recovered population interactions appear on the right-hand sides of Equations 1-3; and, therefore, the SIR models fail to predict the trends in the observed data. This is seen explicitly in Equation 9 which contains the effect of the recovered population that is missing in Equation 3.

### Controlling Epidemics Early

Quantitative mathematical relationships, derived within the FE model *(6)*, characterize the dynamics of an epidemic and illustrate that strong and early intervention is critical. This is highlighted by Equation 46 *(6)* which projects the ultimate number of individuals infected in an epidemic, *N*_∞_, to be exponentially dependent on the number of people a with which a person interacts. The country data provide vivid examples. Both South Korea and New Zealand enacted strong and early interventions compared to other countries (*3, 4*). This is reflected by their *K*_*1*_ values (Table 1). These strong interventions led to earlier peaks in new cases and to far fewer cases than in other countries (Fig. 2): the peak number of new cases in South Korea and New Zealand was 90–99% lower than in other countries, compelling evidence that strong intervention led to exponentially more favorable outcomes.

In a further illustration of the importance of early intervention, actions in the USA began to have an effect around March 26 (Fig. 4B); the number of active cases on that date was 46,442 (Table 1). Using the values of *K*_*1*_ and *K*_*2*_ in Table 1, Equation 46 predicts that the ultimate number of cases should have been approximately 1.4 million. In contrast, had the intervention been implemented and sustained starting on March 10, when there were 62 times fewer (754) cases *(7)*, the model predicts that the ultimate number of cases would also have been 62 times lower, or 23,500. Earlier action could have reduced the ultimate number of projected cases by more than 98%.

The projected estimate of ∼1.4 million total US cases would only have occurred if the effectiveness of the interventions launched on March 26 had been sustained. Unfortunately, a a marked reduction in effective interventions occurred widely in the USA in mid-April, well before the official reopening of the economy *(9)*. This caused a second surge in new cases and a new, third surge has been caused by additional reopening activities in early fall.

The FE model also provides an estimate of the time to the peak of new cases, *t*_max_. Using Equation 54 *(6)* and the values of *K*_*1*_ and *K*_*2*_ from Table 1, the predicted peak in new cases in the USA would have occurred near March 24 if the intervention had begun on March 10. Instead, the USA reached an initial peak on April 11 *(7)*, 18 days later, and it was much higher (Fig. 6B).

Epidemic acceleration, the instantaneous potential to change the pace of the epidemic, can be determined at any point in the epidemic and depends on the social containment actions in effect at that time (Equation 55 *(6)*). Two countries with identical numbers of cases on a given day can, in fact, have different accelerations on the same day, and exhibit different dynamics immediately after that day.

For example, South Korea and New Zealand (Fig. 2) had nearly identical case counts when each imposed strong containment measures (155 cases in South Korea on February 21, and 189 in New Zealand on March 25). Their interventions model as being about equally effective (*K*_*1*_ = 0.195 in South Korea and 0.214 in New Zealand; Table 1). However, since South Korea has a much higher population density than New Zealand (Table 2), it had a much higher number of interactions when the interventions were imposed, and therefore, a higher rate of acceleration. Indeed, the rate of change of new cases <u>was</u> higher in South Korea than in New Zealand; and, the later number of cases in South Korea was higher than New Zealand’s (Fig. 2A, B).

The lessons are clear: early, and strong interventions, especially in countries with indigenously high levels of social interaction, are necessary to stop an epidemic in the initial stages; and reopening actions, enacted too early, can reignite the epidemic dramatically increasing the number of cases. The astonishing magnitude of the effects, driven by only days of delay, derive from the doubly exponential nature of the underlying relationships.

### Ending an Ongoing Epidemic

The FE model can also be used to design measures to end an epidemic in an advanced stage. The management plan is built by first using Equation 12 to predict how many days a given level of intervention, *K*_1_, takes to reduce the new daily cases by a target fraction:

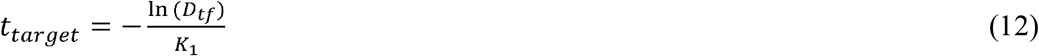

Where *t*_*target*_ is the time to the desired reduction and *D*_*tf*_ is the target as a fraction of the current level of new daily cases.

For example, if a country were to target a 90% reduction of new cases per day (e.g. from 50,000 to 5000 cases per day,), this level can be attained in about 12 days by imposing a containment level of *K*_1_ = 0.2. The New Zealand and South Korea data demonstrate that Equation 12 is valid and that *K*_1_ = 0.2 is achievable for this duration. Both achieved a *K*_1_ = 0.2 for the necessary time which produced a 90% reduction within 13 days (March 2-15, South Korea; April 2-15, New Zealand *(7)*).

In this example, since 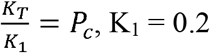 characterizes a lockdown in which people in the country can each only have one plausibly infectious contact with one specific person for the containment duration. This does not mean they cannot contact anyone other than the one person; but they must use care, using masks and proper distancing, to ensure there is no plausibly infectious contact with anyone other than the one person.

Continuing the planning example: Once the initial 90% reduction is achieved, a reasonable next step might be to relax the social containment to a level allowing a viable economy while preventing the epidemic from erupting again. The level of *K*_1_ necessary to achieve a chosen target can be found using Equation 12. If an additional 90% reduction in new cases per day is desired and a period of 90 days is tolerable for that reduction, then the new level of *K*_1_ needed is ∼0.025. This equates to allowing each person to be in contact with 7 specific people, in an infectable way, for 90 days. [This is 3 times less stringent than the original US shutdown level in April, 2020.] Thus, with a well-planned approach, a country can reduce its new daily cases by 99% in approximately 100 days enabling the country to control, and essentially end the epidemic, while maintaining economic viability.

If even *K*_1_ = 0.025 is too restrictive, an alternative, lower *K*_1_ can be chosen, but it must be large enough to avoid a new outbreak A bound for the new value of *K*_1_, low enough to prevent an outbreak and continue to decrease the new cases per day, can be found using Equation 56. Of course, a lower *K*_1_ will extend the time to the additional 90% reduction, but this may be palatable if the initial containment was strong enough.

The progress of interventions is easily monitored using the RCO, as the curve for South Korea illustrates. (Fig. 4A) Had they maintained the implemented level of distancing measures, the data would have followed the initial slope. However, the actual data departed from the slope, heralding the failures in (or relaxations of) social distancing, which were later documented to have occurred during the indicated time frame *(3)* (circled data, Figs. 2F and 4A). Because it summarizes the epidemic dynamics, the RCO can be used to continuously determine if implemented measures are effective or need to be altered.

The RCO, with Equations 12 and 56, form a simple, yet powerful toolbox for policy makers to kraft measures for controlling an epidemic. With only the time series of the case data and a calculator or spreadsheet, specific goals, and the necessary quantitative behavior of the populace to achieve those goals, can be determined.

## Discussion

The data on the current COVID-19 pandemic are widely accessible from a variety of sources, updated daily. The unfolding panorama provides a test bed for models used to predict outcomes and the effects of various interventions. Because different countries have employed different containment strategies *(3-5, 8)*, the world is conducting an epidemiological experiment on a grand scale.

The foundations of current epidemiological modelling are the SIR models *(1)*. In use since 1927, they exist in many variants, both deterministic and stochastic, and their behavior is widely known. It is startling, then, that when the classic SIR model is tested using the currently available data from the COVID-19 pandemic, it fails the most basic test for any model: it projects trends that are the opposite of those easily visible in the data from tens of countries.

Myths die hard. The false “flattening of the curve” myth may well have caused country leaders, especially those most concerned with economic performance, to see social distancing as producing only a modest reduction in the horror of an epidemic peak while significantly prolonging economic disruption. Flattening the curve, as seen in SIR analyses, projects an extension of the epidemic at a high number of cases; however, this notion is completely incorrect

The FE model, a fundamental departure from the SIR models, developed from basic principles and more realistic assumptions, incorporates the effect of the recovered population’s interactions with the susceptible and infected populations and accurately projects the epidemic trends of many countries in a unified way. The model makes clear that short and sharp social distancing produces rapid truncation of epidemic upward trends thereby shortening, not lengthening the time needed to bring the epidemic under control. Second, an indicator of the rate of change in epidemic dynamics (the RCO) allows direct observation of the effectiveness of intervention measures and provides policymakers with an opportunity to react before a new outbreak gains momentum.

Every country and economy can use the FE model to plan and implement the highest level of social distancing measures deemed sustainable to quickly reduce case numbers to levels at which case identification, contract tracing, testing and isolation can be maintained, allowing the return of nearly normal social interactions while minimizing economic consequences.. The ultimate insight from the model is one of hope: the path of an epidemic is not an uncontrollable force of nature nor is epidemic control inevitably the road to economic ruin. Rather, the afflicted population can, through their behavior, choose to control their destiny.

## Data Availability

All data used in this paper are available at the website: https://ourworldindata.org

## Acknowledgments

Drafts of this manuscript were edited by NeuroEdit.

## Funding

This research received no specific funding.

## Author contributions

T. Duclos and T. Reichert: Conceptualization, Writing – Original draft preparation, Writing – Review & Editing. T. Duclos: Methodology, Validation. T. Reichert: Data Investigation.

## Competing interests

Authors declare no competing interests.

## Data and materials availability

The datasets generated and analyzed in the present study are available from the corresponding author on reasonable request.

## Supplementary Materials for

### Materials and Methods

#### The SIR Model

The SIR model, with non-time varying parameters and, is described by the following equations:

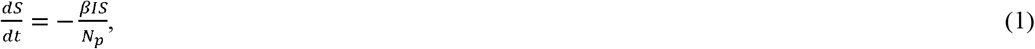

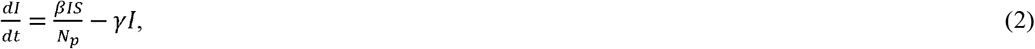

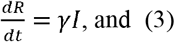

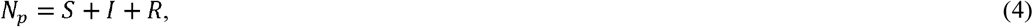

where *S* = number of people susceptible to infection at time *t*; *I* = number of people infected at time *t*; *R* = number of people recovered at time *t*; *N*_*p*_ = total number of people in the population; *β* = rate of contact and transmission; *t*_*r*_ = time of infectiousness; *γ* = rate of recoveries = 1/tr. This formulation of the SIR model is from (1), which also presents a version of the model where *β* and *γ* are functions of time. However, Equations 1–4 retain the general characteristics of the equations with time varying coefficients; therefore, this paper uses Equations 1–4 in the qualitative comparison described in the Main Text.

A solution to Equations 1–4 is easily obtained using straightforward computational methods. Fig. 1 depicts one such solution, computed using a simple Euler method to solve differential Equations 1–4 with a time step of 1 day.

The development of equations 1–4 in (*1*) relied on the basic assumption that the rate of new infections in the susceptible population (*S*) is proportional to the product of the susceptible and infected populations (*I*). On its surface, this assumption seems reasonable. After all, infected people only infect susceptible people. However, it ignores that over time, the population of recovered individuals (*R*) grows and interacts with infected and susceptible people. Since the recovered population grows substantially during the epidemic, interactions between recovered people and the rest of the population significantly alter the epidemic dynamics.

In addition, Equation 3 states that the recovery rate, 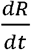, at time *t* is always the same portion of the number of the current infections, *I*(*t*). However, we know that 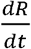 is equal to the number of new infections at time *t* −*t*_*r*_. Therefore, Equation 3 requires that *I*(*t*) also be in constant proportion to the number of new infections at time Since, according to Equation 2, *I*(*t*) grows in a nonlinear fashion with time, Equation 3 is not reasonable.

The neglect of the recovered interactions and the requirement that *I*(*t*) be in constant proportion to the number of new infections at time *t* −*t*_*r*_ in the SIR model raise serious doubts about the validity of the model and suggest we need a more complete model to account for these deficiencies.

#### The FE Model

We begin developing a new model by first separating individuals who were infected into two categories: those who are currently infectious, defined as *I*(*t*), and those who have had the infection, but are no longer contagious, defined as *R*(*t*). The population, *R*(*t*), is also referred to as the recovered population because although they may still be ill, they are no longer infectious. We define people already infected during the epidemic as the sum of these two populations: *N*(*t*) = *R*(*t*) + *I*(*t*).

The goal of developing the new model is to find an expression for *N*(*t*) and for the total number of people who will be infected during the epidemic, defined as co, in terms of the parameters of disease transmission and the behavior of the population enduring the epidemic. To find co, we must first find an expression for *N*(*t*) and then find the limit of *N*(*t*) as time increases. For purposes of this model, we assume that people in the recovered population remain immune after recovery and that the epidemic begins with the introduction of the infection by one individual at time *t=*0; therefore, *I(0) = N(0) = 1*. In addition, we assume that no new infections other than the initial infection are introduced from outside the region of interest during the epidemic.

(Note: Equations 1-12 were introduced in the body of the paper, and the numbering begun in the body has been continued in the supplementary materials. Therefore, some equations in this section may appear to be numbered out of order.)

As we develop the model, we note the following:

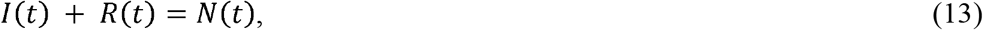

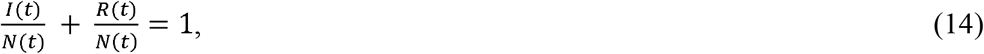

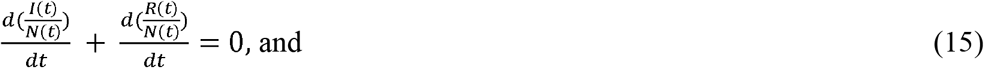

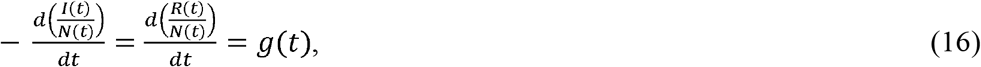

where *g*(*t*)is an, as yet unknown function of time.

We now need to find an expression for the function, *g*(*t*), in terms of the model variables and parameters. We know that 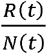 can only change if *I*(*t*) changes, since all the recovered people must have been infected; therefore, 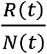 can only change if the quantity 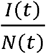 changes.

Thus, we can write the following expression:

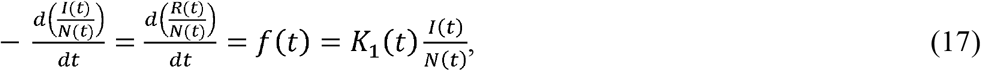

where *K*_1_(*t*)is a function describing the fraction of 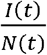 that recovers at time *t* and is initially assumed to be a function of time.

Equation 17 is a simple differential equation in the variable 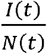, whose solution is

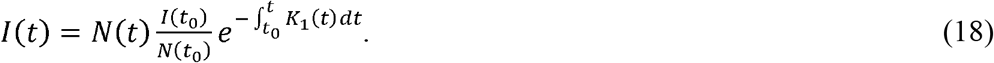

Or, if *K*_1_(*t*) is a constant,

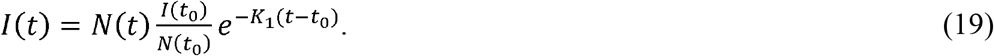

Likewise, the solutions for *R*(*t*) are

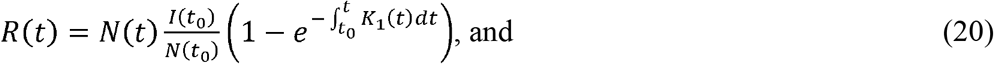

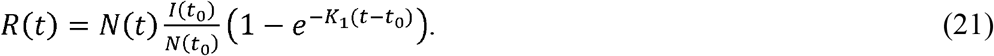

When time *t* = 0, *N*(*t*_0_) = *I*(*t*_0_) = 1 and Equations 18 through 21 become

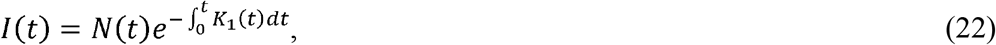

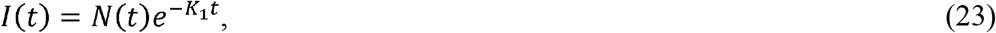

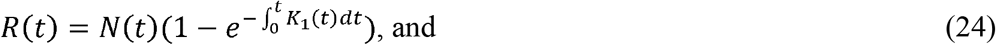

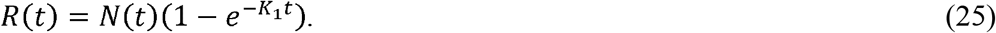

During an epidemic, if every contact made by a person within the population *N*(*t*) could result in an infection, then the disease progression can be represented by the following expression:

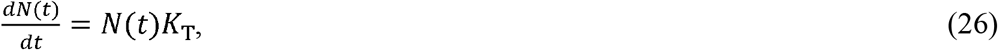

where *K*_T_ is a constant representing disease transmissibility.

However, because *N*(*t*) consists of both infected and recovered individuals, we can rewrite Equation 26 as

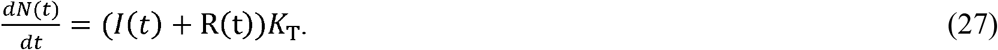

Because population *R*(*t*) cannot transmit the disease, *R*(*t*) *K*_T_ = 0, Equation 27 can be rewritten as

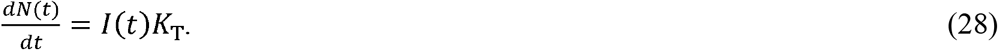

Using Equation 28 and Equations 18 or 19, we can write

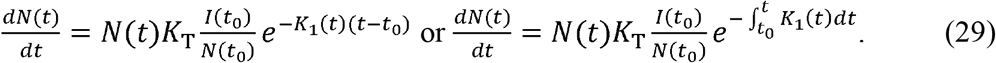

Both forms of Equation 29 can be solved for *N*(*t*), but before we do this, we need to define *K*_1_ more explicitly.

We begin by defining a new parameter, *P*_*c*_, as the average number of specific infectable contacts each individual in the population *N*(*t*) has within the entire population. We use the words, specific infectable, because we mean that each person only interacts with the same people (quantity = *P*_*c*_) in a way they can transmit the disease for the time under consideration. Based on this definition, therefore, the number of interactions between the population *N*(*t*) and the entire population is *N*(*t*)*P*_*c*_ at time *t*. If we also define the rate of contact between *N*(*t*) and the population as *P*_*cr*_, then *P*_*c*_ can be defined mathematically as

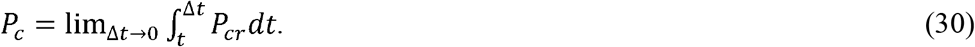

Since interactions among the population during different stages of the epidemic are expected to be relatively stable, we initially assume that the contact rate, *P*_*cr*_, is a constant and therefore *P*_*c*_is also a constant. Additionally, for simplicity in the expressions, we define *F*_*i*_(*t*) as the fraction of *N*(*t*) that is infected:

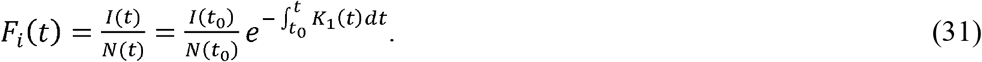

Using these definitions, we can write the following expression using Equation 29:

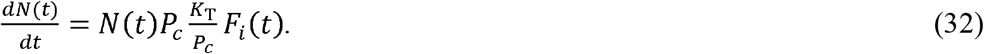

Equation 32 can then be written as the following difference equation:

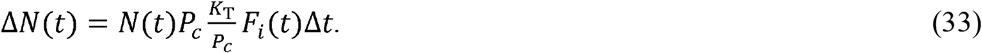

Equation 33 can also be rewritten in the following manner:

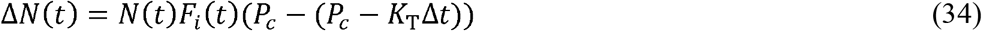

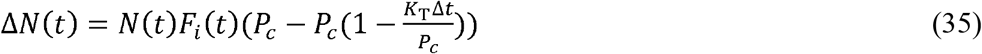

In Equation 34, the term (*P*_*c*_ − *K*_T_Δ*t*) is the change in infectable contacts in the time Δ*t* due to infections that occurred during time Δ*t*. Likewise, the term 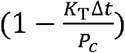 in Equation 35 is the change in the fraction of infectable contacts during Δ*t*. Since *P*_*c*_is independent of the infectable contacts, the change in the fraction of infectable contacts must be the fraction that *F*_*i*_(*t*) changes during Δ*t*. Rewriting Equation 35, we can express this mathematically as

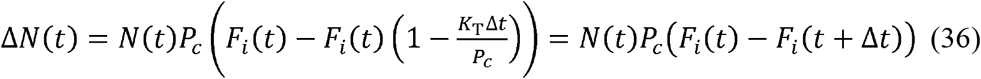

We can now use Equation 36 to develop a recurrence relationship to explain the nature of *K*_1_

At *t* = 0, *F*_*i*_(0) = 1 and Equation 36 becomes

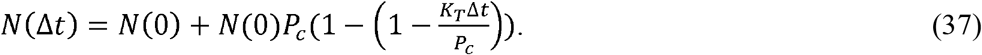

We should also note that

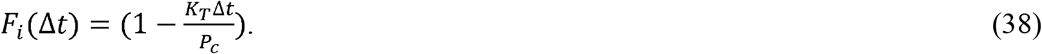

In the next time step, applying Equation 35 to Equation 37, we find the following expression:

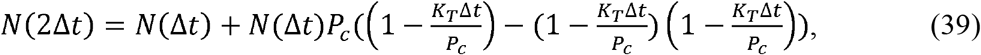

noting that 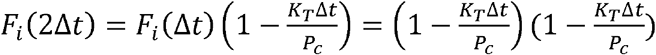.

Equation 39 can be simplified to

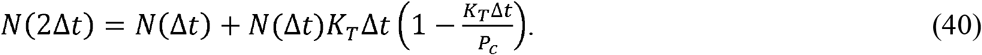

Subsequent application of the same logic leads to the recurrence relationship

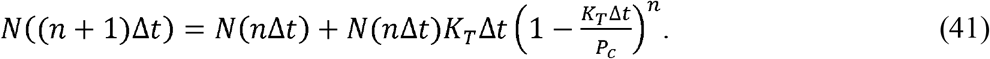

Rearranging terms and defining *n*Δ*t* = *t*, we can write the following:

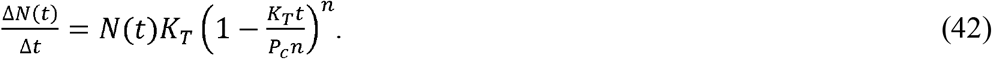

If we now allow Δ*t* →0 and therefore, *n*→∞, and also note that 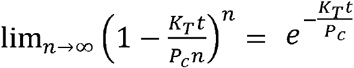, we get the following expression for 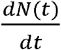:

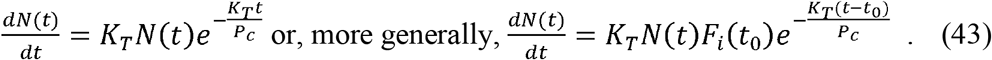

Comparing Equation 43 to Equation 29, we can see that 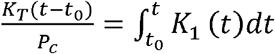 or if *P*_*c*_ is a constant, 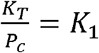 or 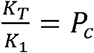.

Returning to *P*_*c*_, we will now further explain this parameter. As defined, *P*_*c*_ is the number of specific infectable contacts a member of the population *N*(t) has within the entire population and is a function of the population’s behavior. Initially, we assume this to be a function of population density and that people cover a constant average effective area per unit of time. We define this area as the effective area rate, *A* _1*r*_. This quantity is assumed to be a constant because the fraction of the population an average person interacts with on any given day presumably remains stable for a given time period.

Using these assumptions, we can write an expression for *P*_*cr*_:

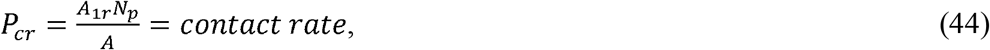

where *N* _*p*_ = the population of the region with the infection, *A* = the area of the region, and 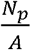 = the population density.

*P*_*cr*_ is proportional to both the population’s behavior, *A* _1*r*_, and the population density, 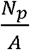. Since both the population’s behaviour and the population density represented by *A* _1*r*_ and 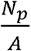 are likely to remain constant for many days running, for the initial model development, we can also assume *P*_*cr*_ is constant and, therefore, *P*_*c*_ is constant. Additionally, *K*_T_ is a constant, therefore *K*_1_ is constant for the duration that *P*_*c*_ remains constant. Of course, we only expect this to be true on a piecewise basis because, eventually, in response to the epidemic, populations resort to measures such as social distancing, quarantines, and re-openings that significantly change the contact rate, and therefore *P*_*cr*_. The piecewise variance of is *K*_1_ addressed later in the this section.

Similar to the way we defined *P*_*c*_ using *P*_*cr*_, we can define a quantity *A*_1_ in terms of *A*_1*r*_: 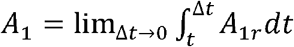, where *A* _1_ is the effective specific area traversed by a person. In this case, “specific” has the same meaning as it has for *P*_*c*_. That is, each person only traverses the same area for the duration of the time under consideration. We also call *A* _1_ the “effective area” because the population will not be evenly dispersed throughout a given region within a country^10^.

If we take this into account, then

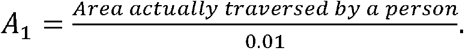

From the preceding discussion, we can now write an expression for *K*_1_(*t*):

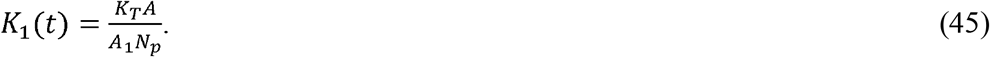

Equation 45 shows that, under the initial assumptions of immunity and no cases from outside, *K*_1_> 0. Also, since *K*_1_ is inversely proportional to *A* _1_, as people reduce the area they traverse per unit time by increasing social distancing or taking other measures, *K*_1_ gets larger due to the lowered contact rate. In other words, *K*_1_ is inversely proportional to the strength of social distancing interventions implemented during an epidemic.

Having defined *K*_1_ as a piecewise constant, we can now complete the derivation of the model. Using the expression for 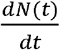 from Equation 29, we arrive at Equations 5 and 6 (presented in the Main Text):

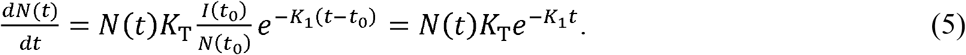

Equation 5 can also be rewritten as

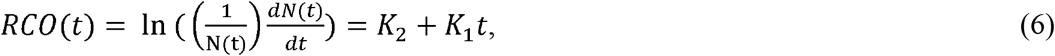

where 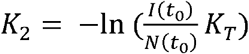 and *t*_0_ is the time between the start of the epidemic and the time at which the first data point is measured in the epidemic. Equation 6 is the equation that was fit to the country data in Figures 2 and 4, used in the RCO projections of Figures 1 and 3, and to find the parameters in Table 1.

The expression ln 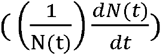 is a convenient operator in that it transforms Equation 5 into a linear equation. We call this expression, ln 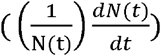, the rate of change operator (RCO) and, by its definition, it is related to the rate of change of new cases. Note that if *K*_1_(*t* − *t*_0_) were ever to approach zero, the growth of the epidemic would be nearly exponential.

Using Equation 19, we can show that 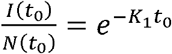. Substituting this relationship for 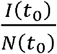 into Equation 5 and then integrating Equation 5, we arrive at Equation 7, which is the expression for the total number of infections, *N*(*t*):

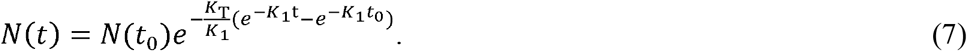

Taking the limit of Equation 7 as *t* → ∞, we can also obtain the following expression for the total number of individuals who will be infected by the epidemic:

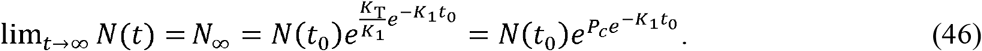

Equation 46 introduces an important and rather abstract concept, not previously discussed. Mathematically, it demonstrates that the number of infections to be expected, *N*∞, and the behavior of the population, *P*_*c*_, are interdependent in the sense that changes in *P*_*c*_ have an exponential effect on the final number of infections that will come to be. This means that small changes in population behavior dramatically affect the epidemic’s outcome. It also means that the eventual number of cases produced by the epidemic is not foreordained; but rather a strong function of interventions introduced.

Additional insight into the model is gained from the following relationship, derived from Equations 5, 7, and 46:

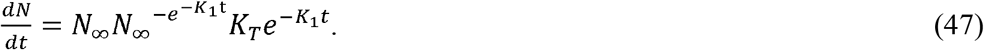

In words, the form of Equation 47 is

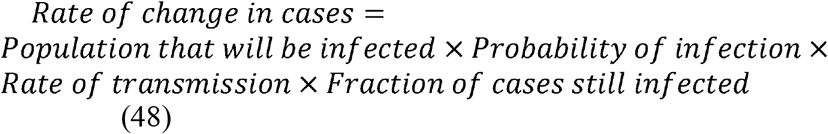

or

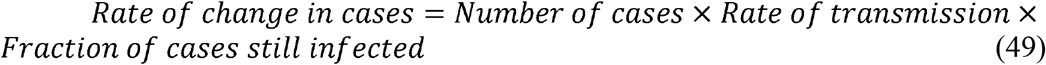

Equations 48 and 49 illustrate the logic of the FE model in terms of probabilities.

Using the prior definition *F*_*i*_ = *Fractions of cases still infected*, and if *t*_0_ = 0, we can use Equation 7 to write this simple expression for the model:

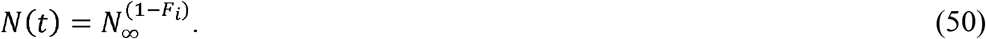

#### The FE Model in Terms of S, I, and R

We can write expressions for *I(t)* and *R(t)* as

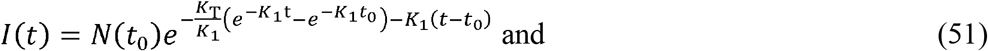

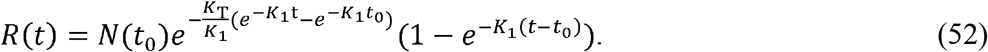

If we define the susceptible population during the epidemic as the number of people who will eventually become infected and call this population *S*(*t*), we obtain the following expression:

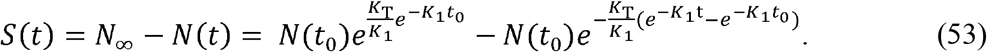

Equation 53 shows that the susceptible population is also not a pre-ordained constant; rather, the number of susceptible people in a population changes as the behavior of the population changes. As 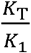 increases or decreases with any change in *P*_*c*_, *S*(*t*) will also change, regardless of the change in infections.

We can rewrite the differential equations of the FE model cast in SIR variables as

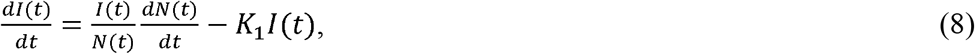

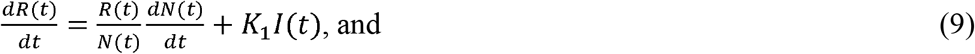

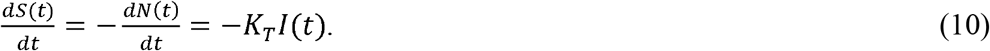

We gain additional insight as to the meaning of these equations by looking at Equation 8 in more detail. Equation 28 can be used to rewrite Equation 8 as

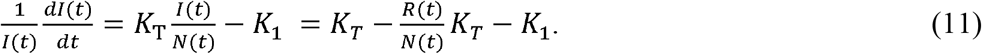

The left-hand side of Equation 11 is the ratio of the rate of change in the number of new infections per person currently infected. The first two terms on the furthest right-hand side of Equation 11, 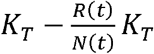, describe the rate of successful infection. These terms account for contacts by the recovered population in a way that SIR models fail to. Furthermore, since *K*_T_ is the rate of infections an infected person causes per infectable contact, the terms 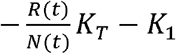 must represent the rate of recovery per infected person, which leads directly to Equation 9.

The FE model is completely consistent with Equations 4, 14, and 15. This is evident when adding together appropriate combinations of Equations 8 through 10.

#### Additional properties of the FE model

If we differentiate both sides of Equation 5, we can derive four important expressions that describe the dynamics of an epidemic. The first is

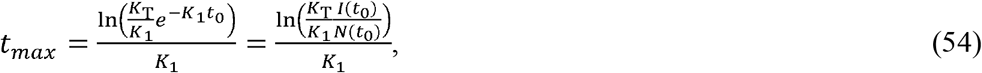

where *t*_*max*_ = the time to the peak of new infections.

Equation 54 demonstrates the relationship between the strength of social intervention measures, 1, and the time to the peak of new infections. When the social interventions are stronger (larger *K*_1_), the time to the peak will be shorter.

The second important expression is the rate of acceleration of the epidemic:

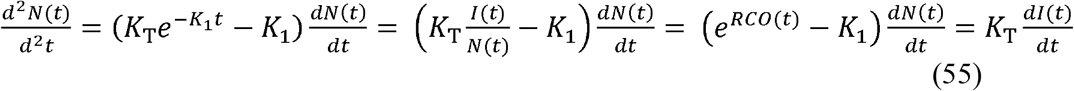

We can use Equation 55 to compare acceleration in the early stage of the epidemic between two countries with different population densities. Equation 55 can also be used to judge whether the control measures in place, represented by 1, are effective enough. If the value of the term 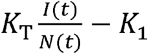 is positive, then the control measures are not strong enough. Conversely, when this term becomes negative, the epidemic is being brought under control.

The furthest right-hand portion of Equation 55, 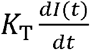, can be interpreted intuitively. The epidemic acceleration is linearly related to the rate of change in the infections. That is, when there are more daily new infections than daily recoveries, the epidemic is accelerating and expanding. Conversely, when there are fewer daily new infections than daily recoveries, the epidemic is slowing and will end if the containment measures are kept in place long enough to extinguish the outbreak. This leads to the inescapable conclusion that the goal of all containment measures must be to limit the creation of new infections as fast as possible to increase the rate of deceleration to the highest level possible.

The minimum value of *K*_1_ that will begin to bring the new cases per day down occurs when the acceleration is less than zero. If we set the left-hand side of Equation 55 to zero and solve for

*K*_1_, we arrive at the relationship for this value of *K*_1_:

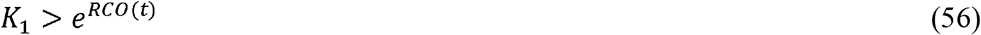

Since the value of the *RCO*(*t*) is always measurable during the course of the pandemic, the minimum value of *K*_1_and therefore, *P*_*c*_, to reduce the number of daily cases can be determined at all times. This information can be used to quickly determine what level of infectable social contact is allowable to continue to decrease the number of new daily cases.

A fourth, and perhaps most important, relationship can be derived from Equation 5. In Equation 5, the term, 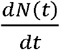, is the rate of new cases. In the figures, this is the new cases per day.

If we define a desired target for the new cases per day at a future time, *t*_*target*_, then we can define a new quantity, *D*_*tf*_, the desired fraction of the current new cases as:

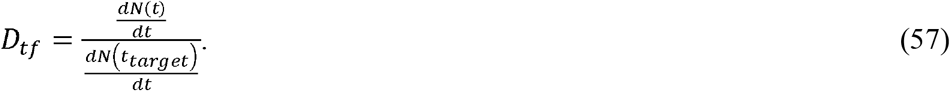

Using Equations 5 and 7, we arrive at the following expression:

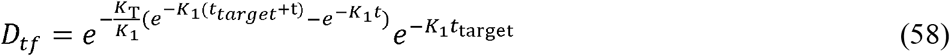

If *t* ≫ *t*_*target*_, then 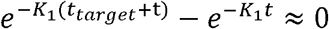 and we can derive Equation 12 from the remaining terms:

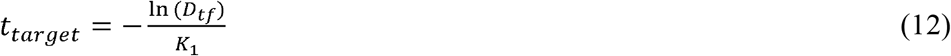

Equation 12 quantitates the number of days, *t*_*target*_ a level of social containment, *K*_1_, will be necessary to achieve a fraction, *D*_*tf*_, of daily cases compared to the current level.

#### Piecewise Time-Varying Parameters

Thus far in the development of the FE model, *K*_1_ has been assumed to be constant over time. The model is readily extended to allow *K*_1_ to vary with time in a piecewise manner. Here, we develop equations describing the analogous model in which *K*_1_ is constant in finite intervals of time, between which it changes. This addresses scenarios in which the population interactions change at particular points in time, but remain constant, or nearly so, in the intervals between those changes. In addition, because this is a common happenstance, we develop an expression for *K*_2_for use in Equation 7 when the model is fit to data that does not start at time *t* = 0. If *K*_1_ and *K*_2_are functions of time, Equation 5 can be rewritten as

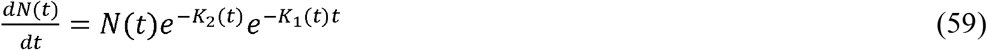

and Equation 7 as

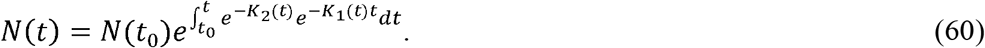

Distancing measures tend to be constant for many days at a time. Thus, for this analysis, we assume *K*_1_ is piecewise linear. Therefore, we can calculate *K*_2_ for any time, *t*_*n*_ when *K*_1_ changes:

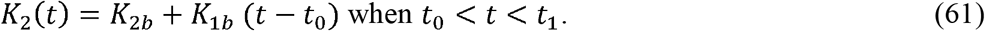

If *t*_0_ = 0, then *K*_1*b*_ and *K*_2*b*_ are the baseline levels of these parameters, and 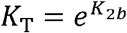. These three *K* values represent the epidemic dynamics during the initial stages, before any containment measures are implemented.

As time passes, *K*_2_ becomes

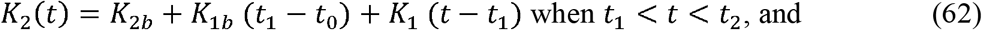

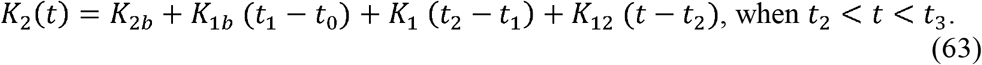

Noting the pattern as t increase we can rewrite Equations 62 and 63 as

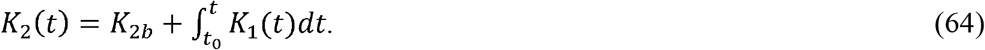

For any interval, *t*_*n*_ < *t* < *t*_*n*+1_where *K*_1_ is a constant, *K*_2_ (*t*) can be expressed as

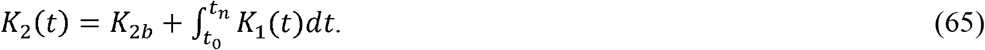

Equations 5 and 7 can also be rewritten as

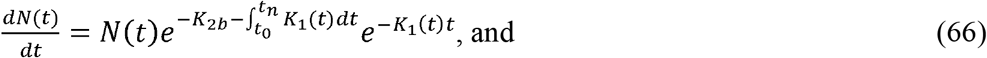

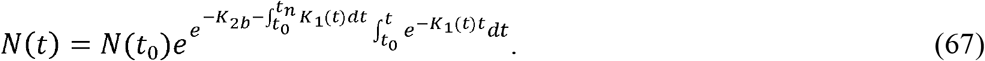

An expression for what happens in an epidemic, *before* and *after* social distancing measures are implemented, can be especially useful. A typical, perhaps worst-case, scenario might be “reopening”, in which social distancing measures are withdrawn and social intercourse returns to normal levels. In that instance, *K*_1_ (*t*) becomes *K*_1*b*_ (*t*) at time, *t*_n_, when distancing measures are lifted. Using Equation 5, we arrive at the following expressions:

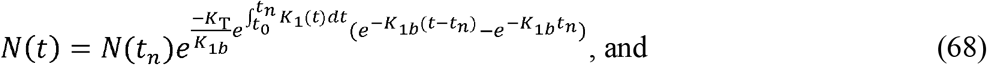

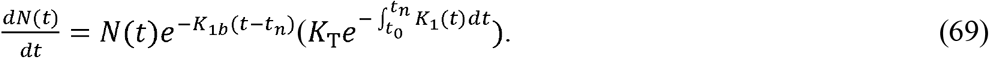

Equations 68 and 69 predict the epidemic’s progression before and after interventions are instituted. They demonstrate that the dynamics of the epidemic will depend on prior containment measures, as shown by the appearance of the expression 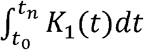 in both equations. This means that as long as the initial assumptions of immunity remain valid and no new infections are introduced from outside the area, then when interventions are relaxed, outbreaks can still grow nearly exponentially, but the growth of second and subsequent waves will not grow as fast as the initial outbreak. This is a consequence of the fact that *K*_1_ > 0 at all times. Since *K*_1_ is proportional to the inverse of the effective area traversed by an individual, it is proportional to the number of social interactions, and social interactions can never be less than zero.

If the immunity wanes or new infection from the outside are introduced to a previously uninfected area, then *K*_1_ may become less than zero for a limited period of time. If, in fact, *K*_1_ does become less than zero, this indicates that either immunity is waning in the population or that new infections have come to a previously unaffected area and a major outbreak is occurring.

The new outbreak can be modelled using Equation 7 or 67 by adding another term to the equation, using a time offset to the time of the outbreak start and choosing an appropriate *K*_1_ to represent the behavior of the population in the outbreak area. Any number of these terms can be added to model multiple outbreaks. Regardless of how the outbreak is modelled, a negative *K*_1_ requires immediate action, within days, from policy makers to strengthen intervention measures and prevent the outbreak from overwhelming prior progress in controlling the epidemic.

#### Finding Model Parameters from Country Data

When Equation 45 is substituted into Equation 7 and solved form *K*_T_*t*, we find the following expression:

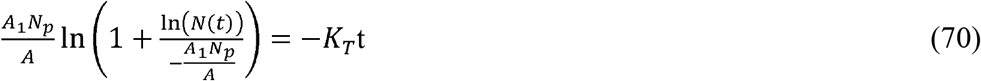

If we define 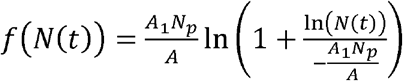, then we can also write this expression:

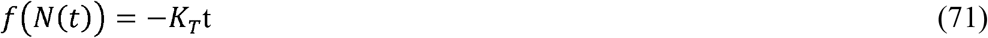

If *K*_T_ is a constant, then Equation 71 predicts that *f*(*N*(*t*)) is a linear function of time. All the quantities on the left-hand side of Equation 70 can be derived from the country data, with the exception of 1. Therefore, we can estimate the value of *K*_T_ using Equation 70 (or 71) to find the value of *A* _1_ that best fits a straight line.

## Notes

### Competing Interest Statement

The authors have declared no competing interest.

### Funding Statement

No external funding was received

### Author Declarations

This is not applicable to this paper

